# Patterns and Predictors of Weight Loss with Exenatide Treatment in Overweight and Obese Women

**DOI:** 10.1101/2020.06.11.20128645

**Authors:** Jody Dushay, Eleftheria Maratos Flier, Robert E Gerzsten, Megan Rodgers, Brent Heineman, Alexandra Migdal, Natasha Kasid, Tahereh Ghorbani, Elena Toschi, Juliet Tripaldi, Anjali Nath, Zsu Zsu Chen, Minh Phan, Long Ngo

**Affiliations:** Beth Israel Deaconess Medical Center, Boston MA

## Abstract

In the medical management of obesity, treating physicians observe significant heterogeneity in responses to pharmacotherapy. Indeed one of the most important clinical questions in obesity medicine is whether we can predict how an individual will respond to a particular pharmacotherapeutic agent. The present study examines patterns and predictors of weight loss among overweight and obese women who demonstrated early robust response to twice daily exenatide treatment.

182 women were assigned using single-blind randomization to either treatment with twice daily exenatide injections or to matched placebo injections with dietary counseling. Women who demonstrated > 5% weight loss after 12 weeks of treatment were deemed high responders and remained on study treatment for up to 52 weeks; women who lost < 5% body weight at 12 weeks were deemed low responders and stopped study treatment. We additionally characterized individuals who lost > 10% of body weight as super responders. Our primary outcome was change in body weight; secondary outcomes included changes in metabolic parameters including lipids, waist circumference, resting energy expenditure, and response to a meal tolerance test. We also performed an exploratory metabolomic analysis.

Consistent with published literature, we observed individual heterogeneity in the weight loss response to exenatide and diet/placebo. Although there was no significant difference between treatment groups in the percentage of participants who achieved > 5% weight loss (56% of exenatide group and 76% of diet/placebo group), or those who achieved > 10% weight loss (43% of exenatide group and 55% of diet/placebo group), in both cases there was a trend toward a higher response rate in the group that received placebo with dietary counseling. In addition to achieving similar average weight loss, both treatment groups also demonstrated similar maximum weight loss. The range of maximum weight loss was greater in the diet/placebo group and there was more weight regain among individuals in the exenatide group compared to the diet/placebo group. In our exploratory metabolomic analysis, we observed lower baseline circulating cysteine concentrations in the exenatide responder group and we also found a trend toward higher baseline levels of serotonin, aminoisobutyric acid, anandamide, and sarcosine in the exenatide super responder group. We did not identify any metabolic predictors of weight loss in either the exenatide or the diet/placebo treatment group.

## Background

The growing obesity epidemic has led to an urgent need for effective treatment options. Recent figures from the Center for Disease Control report the prevalence of obesity among adults in the US at 39.8% in 2017^1^. While individuals with obesity can achieve robust weight loss with structured interventional and behavioral programs, weight regain is very common^2,3^. A recent meta-analysis showed that people who lost weight at an average of 9.9kg through diet alone, or through a combined diet and exercise regimen, regained approximately 50% of the weight within one year^4^. Treatment of obesity with bariatric surgery is often associated with significant long-term weight loss, however many people are not eligible for surgery due to not meeting strict NIH criteria^5^, lack of insurance coverage, or comorbidities. Additionally, many who qualify for bariatric surgery are not willing to pursue surgical weight loss for a variety of reasons^6,7^. Investigating pharmacologic agents, including factors that contribute to heterogeneity in response to medical treatment, is an area of high clinical and public health significance.

One pharmaceutical class generating great interest in this field is the glucagon like peptide 1 receptor (GLP-1R) agents, including exenatide, liraglutide, dulaglutide and semaglutide. Glucagon like peptide 1 is a peptide hormone secreted from the enteroendocrine L cells in the gut. The first approved GLP-1R agent, exenatide, was developed for the treatment of type 2 diabetes, as the primary actions of GLP-1 are to stimulate pancreatic beta cells to increase insulin and to decrease glucagon secretion^8^. In individuals without diabetes, exenatide, liraglutide and semaglutide have been shown to cause significant weight loss and improvement in metabolic parameters^9–12^. A recent review article noted that treatment with liraglutide, the first GLP1R agent approved by the FDA specifically for weight loss, can lead to 5-10% reduction of body weight ^9–11^. Semaglutide, the most recently approved GLP-1R agent, has been shown to be associated with even more weight loss, up to 14% at higher doses, in individuals with obesity without diabetes^13^.

We have previously reported variable weight loss in women with overweight or obesity and without diabetes treated with twice-daily exenatide for 16 weeks^12^. It remains unknown why this heterogeneity in response to pharmacotherapy exists. Early weight loss of >4% body weight with liraglutide has been shown to predict 1-year weight loss, but additional factors that predict or account for variability of weight loss achieved with GLP-1R agents are not well understood^13,14^. The present study examines patterns and predictors of long-term weight loss among women with overweight and obesity who demonstrate initial robust weight loss with twice-daily exenatide treatment.

## Methods

All study procedures were approved by the Beth Israel Deaconess Medical Center (BIDMC) institutional review board (ClinicalTrials.gov Identifier: NCT01590433). All study visits were conducted at the Harvard Catalyst Clinical Research Center at BIDMC in Boston, MA in accordance with the Declaration of Helsinki.

Two hundred and forty-nine women were screened and 182 enrolled in a 52 week single-blind interventional trial (*Figure S1)*. Participants were randomized to treatment with either exenatide (n=127) or placebo with diet intervention (n=55, hereafter referred to as the diet group). Eligibility criteria included age 18-70, BMI 25.0 kg/m^2^ - 48 kg/m^2^ and no acute or poorly controlled chronic medical problems. Pregnancy, lactation, history of pancreatitis, and previous use of exenatide were major exclusion criteria. All subjects administered twice daily subcutaneous injections of either exenatide or matched placebo. Subjects in the diet group received nutrition counseling at every study visit, which included recommendations for a hypocaloric diet (500 kcal/day reduction, with total caloric requirement assessed from measured resting metabolic rate). The exenatide group did not receive any additional dietary counseling. Study visits took place every 2 weeks for the first month and then every 4 weeks for the duration of the study. Subjects were included in the data analysis if they completed at least 12 weeks of treatment (exenatide n=75, diet n=33).

At 12 weeks, subjects who had lost ≥ 5% of their body weight were categorized as high responders and continued participation in the study for up to 52 weeks. Those who did not achieve 5% weight loss were categorized as low responders and their participation ended. Additionally, participants who lost ≥10% body weight at any point during the study were classified as “super high responders.”

Body weight, waist circumference, vital signs and body composition (measured using bioelectrical impedance analysis) were assessed at each visit. Comprehensive metabolic visits occurred after an overnight fast at weeks 0, 12, 24, and 52 and included a blood draw for hematology, electrolyte, and lipid panels; measurement of resting energy expenditure using the Sensormedics Vmax Encore 29 indirect calorimeter (CareFusion,Respiratory Care Inc.,Yorba Linda, CA); and measurement of the thermic effect of food with a liquid meal tolerance test (Boost, 253 kcal, 6.3g fat, 34.8g carbohydrate, 15.8g protein). Blood was drawn 0, 1, 2, 3 and 4 hours following ingestion of Boost for measurement of postprandial triglyceride and insulin levels.

In an exploratory analysis, we sought to identify metabolites that were associated with treatment response by leveraging a targeted liquid chromatography-mass spectrometry (LC-MS) method that measures ∼150 known metabolites^15^. This platform measures polar metabolites that fall into 8 classes: 1) amines; 2) amino acids and amino acid conjugates; 3) bile acids; 4) sugars and sugar phosphates; 5) indoles and indole derivatives; 6) organic acids; 7) purines and pyrimidines; and 8) acylcarnitines. In addition, the platform measures a total of 19 acylcarnitines spanning distinct fatty acid species that contain short-, medium-, and long-carbon chains. We measured these metabolites in a subset of subjects who received exenatide (36 high responders and 28 low responders) and a subset of diet responders (n=23). Metabolomic analysis was done using hydrophilic interaction liquid chromatography (HILIC) coupled with multiple reaction monitoring-based mass spectrometry to measure ∼150 endogenous metabolites. Metabolites were extracted from 10 µl of plasma using acetonitrile and methanol. Samples were spiked with deuterated internal standards for quality control. The extracts were separated using reverse-phase chromatography and detected by a coupled 4000 QTRAP mass spectrometer in positive mode (Sciex). In addition to sample injections, control pooled plasma samples were included and spaced every 10 sample injections to gauge the effectiveness of normalization, to adjust for temporal drift of the instrument, and to calculate the coefficient of variation for each metabolite. Metabolite quantification was determined by integrating peak areas using MultiQuant software (Sciex).

## Data Analysis

When observing the p-values associated with various variables and group labels, chi-squared modeling and t-tests were conducted to look at parametric testing and non-parametric testing. As the data did not always follow a normal distribution, both methodologies were used to compare the splay of that data, with the reporting value being taken from the chi-squared test.

The data used to compare exenatide and diet high responders were taken longitudinally from baseline until the cut-off visit at week 12. Groups were then compared to determine if there were any significant parameters that could predict high responder or super responder status within the treatment groups. The Kruskal Wallis test was used in the models investigating predictors of weight loss in either treatment group. Results are reported as adjusted odds ratios.

To assess the potential association between weight change (percent change from baseline to 3 months of treatment) and the potential predictors of weight loss (age, weight, total cholesterol, waist circumference and resting energy expenditure by treatment group), we used multivariable logistic regression.

For the metabolomic data analysis, mean CVs for analytes measured were ≤15%, and closer to 6% for abundant analytes such as amino acids. Metabolites with CVs ≥ 30% were excluded from analysis^16^. Ninety-two baseline fasting plasma samples were studied from the two treatment groups (exenatide high and low responders n = 64, diet/placebo n = 23). Metabolite concentrations determined by LC-MS peak intensities that were normalized using log transformation. A majority of metabolite concentrations were successfully normalized, however, several continued to have non-normal peak distributions as determined by the Shapiro-Wilk test. Wilcoxon-rank sum tests were used to compare baseline metabolite concentrations between responders in the exenatide and diet treatment groups as well as responders verses non-responders, and super-responders verses responders within each treatment group. (www.r-project.org).

## Results

Demographics of the study population are summarized in Table 1. There were no significant differences in baseline parameters between the exenatide and the diet groups. There were similarly no differences in baseline characteristics within the responder population (Table S1).

**Table 1:** Baseline characteristics of the study population. Values are shown as mean ± SD.

|  | Exenatide (n=75) | Diet (n=33) | *p* |
| --- | --- | --- | --- |
| Age (years) | 43.9 ± 11.9 | 44.6 ± 13.7 | 0.78 |
| BMI (kg/m<sup>2</sup>) | 36.1 ± 5.3 | 34.8 ± 5.0 | 0.21 |
| Weight (kg) | 95.9 ± 14.6 | 94.0 ± 13.1 | 0.51 |
| Systolic BP (mmHg) | 125.1 ± 12.6 | 121.2 ± 14.4 | 0.16 |
| Diastolic BP (mmHg) | 73.6 ± 9.3 | 71.8 ± 12.0 | 0.40 |
| Heart Rate (bpm) | 67.9 ± 10.6 | 66.1 ± 10.8 | 0.41 |
| Total Cholesterol (mg/dL) | 193.2 ± 36.1 | 194.7 ± 32.9 | 0.84 |
| HDL Cholesterol (mg/dL) | 59.6 ± 16.8 | 60.3 ± 11.0 | 0.83 |
| LDL Cholesterol (mg/dL) | 113.0 ± 31.0 | 109.6 ± 29.8 | 0.61 |
| Triglycerides (mg/dL) | 105.7 ± 52.2 | 109.4 ± 54.1 | 0.74 |
| Hemoglobin A1c (%) | 5.6 ± 0.3 | 5.6 ± 0.3 | 0.33 |
| Waist Circumference (cm) | 112.1 ± 17.5 | 110.7 ± 11.9 | 0.70 |
| Resting Energy Expenditure (kCal/d) | 1552.1 ± 202.6 | 1522.9 ± 199.9 | 0.49 |
| Percent Body Fat (%) | 43.6 ± 4.9 | 43.7 ± 4.1 | 0.84 |

### Weight Loss and Regain Among High Responders and Super High Responders

In total, 56% percent of the exenatide group and 76% of the diet group were categorized as either high responders or super responders (Figure 1A), however this difference was not statistically significant. Individual weight loss trajectories were highly variable in both exenatide and diet high responder groups (Figure 1B and 1C). Figure 2 shows individual and average weight loss among the high and low responder groups over the duration of the treatment period for each group. After 12 weeks of treatment, all groups except diet nonresponder had significant weight loss (Figure 2, table 2). Subjects in the exenatide high responder group had an average weight reduction of 6.2kg and BMI reduction of 2.3, while exenatide low responders achieved an average weight reduction of 1.9 kg and BMI reduction of 0.7 (Table 2). Treatment with diet and placebo injection was associated with a 7.2kg reduction in weight and a BMI reduction of 2.9 in the high responder group, while those in the low responder group had a nonsignificant increase in weight (+ 0.6kg) and BMI (+ 0.2, Table 2).

**Table 2.**
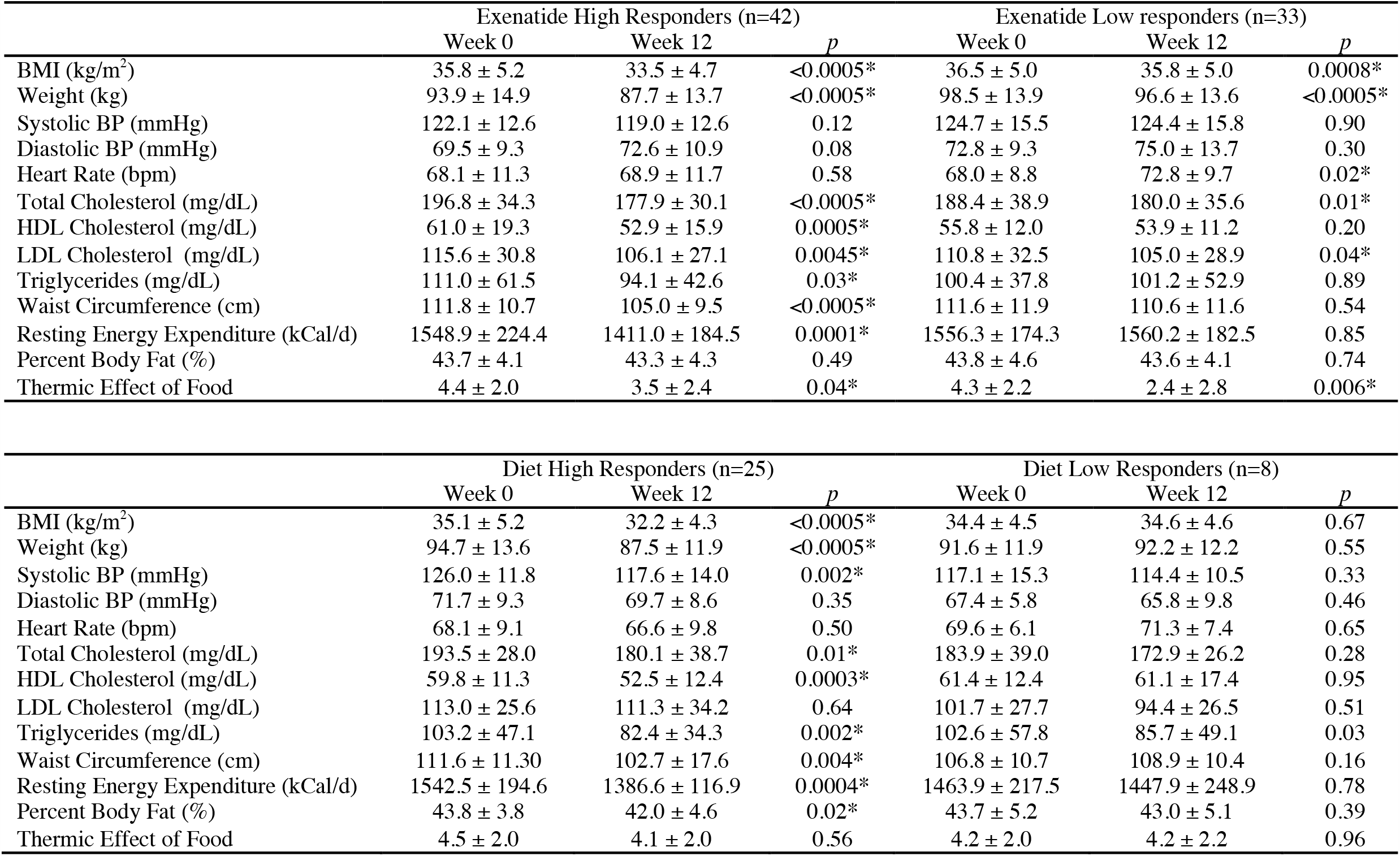
Characteristics of all four study groups before and after twelve weeks of treatment with either exanetide or diet. Values are shown as mean ± SD. P values < 0.05 were considered significant.

**Figure 1:**
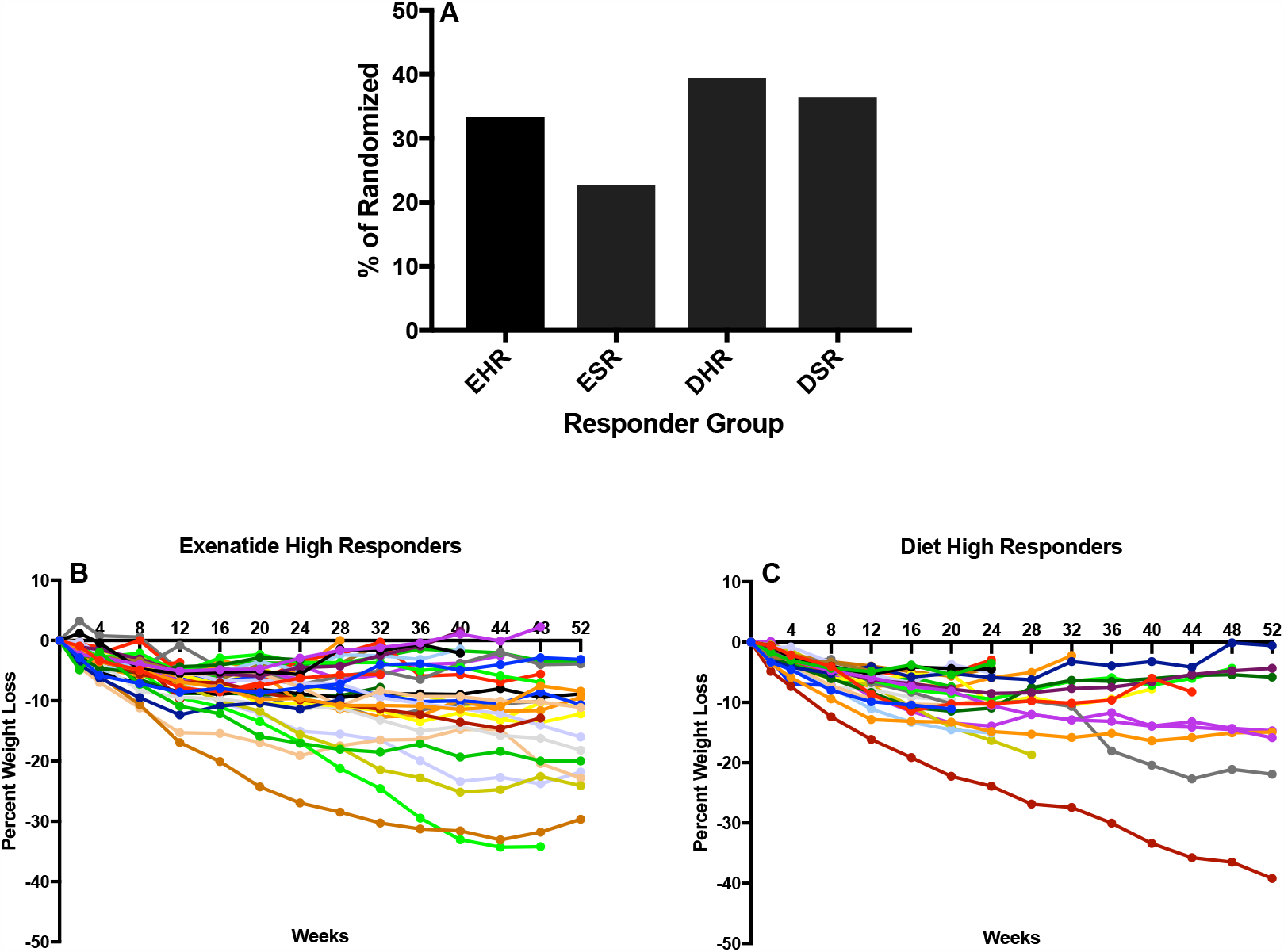
(A) Percentage of high and super responders among those randomized to exenatide and diet groups. (B-C) Individual weight trajectories in exenatide high responder group (B) and diet high responder group (C). EHR exenatide high responder. ESR exenatide super responder. DHR diet high responder. DSR diet super responder.

**Figure 2:**
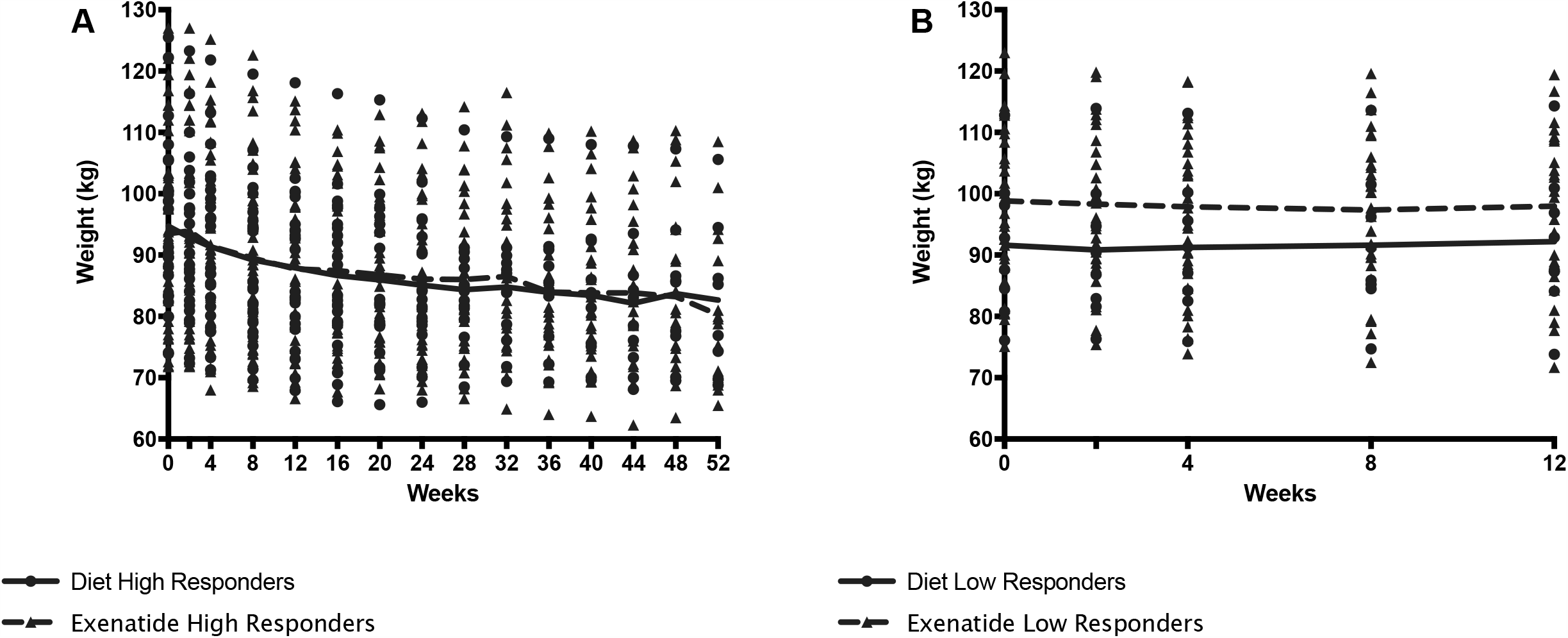
(A). Weight loss among exenatide and diet high responders over 52 weeks (B) Weight loss among exenatide and diet low responders over 12 weeks. By protocol design, low responders ended study participation at 12 weeks.

Maximum (kilograms) and percentage weight loss were similar between exenatide and diet high responders (exenatide 11.65 kg, 6.8%, diet/placebo 11.91 kg, 6.9% Figure 2a and 2c), however the range of maximum weight loss was larger in the diet high responder group (2.90kg – 47.90kg) compared to the exenatide high responder group (4.20kg – 39.20kg). Super responders lost significantly more weight than high responders both treatment groups (Figure 3A). We observed weight regain during the treatment period. Figure 3 shows that the majority of participants experienced weight regain at some point during the study treatment period (Figure 3B-C). The average weight regain during the treatment period in the exenatide high responder group was higher than the regain in the diet high responder group, however this difference was not significant (2.8 +/- 2.7kg vs 2.1 +/-2.0kg, Figure 3B, 3C).

**Figure 3:**
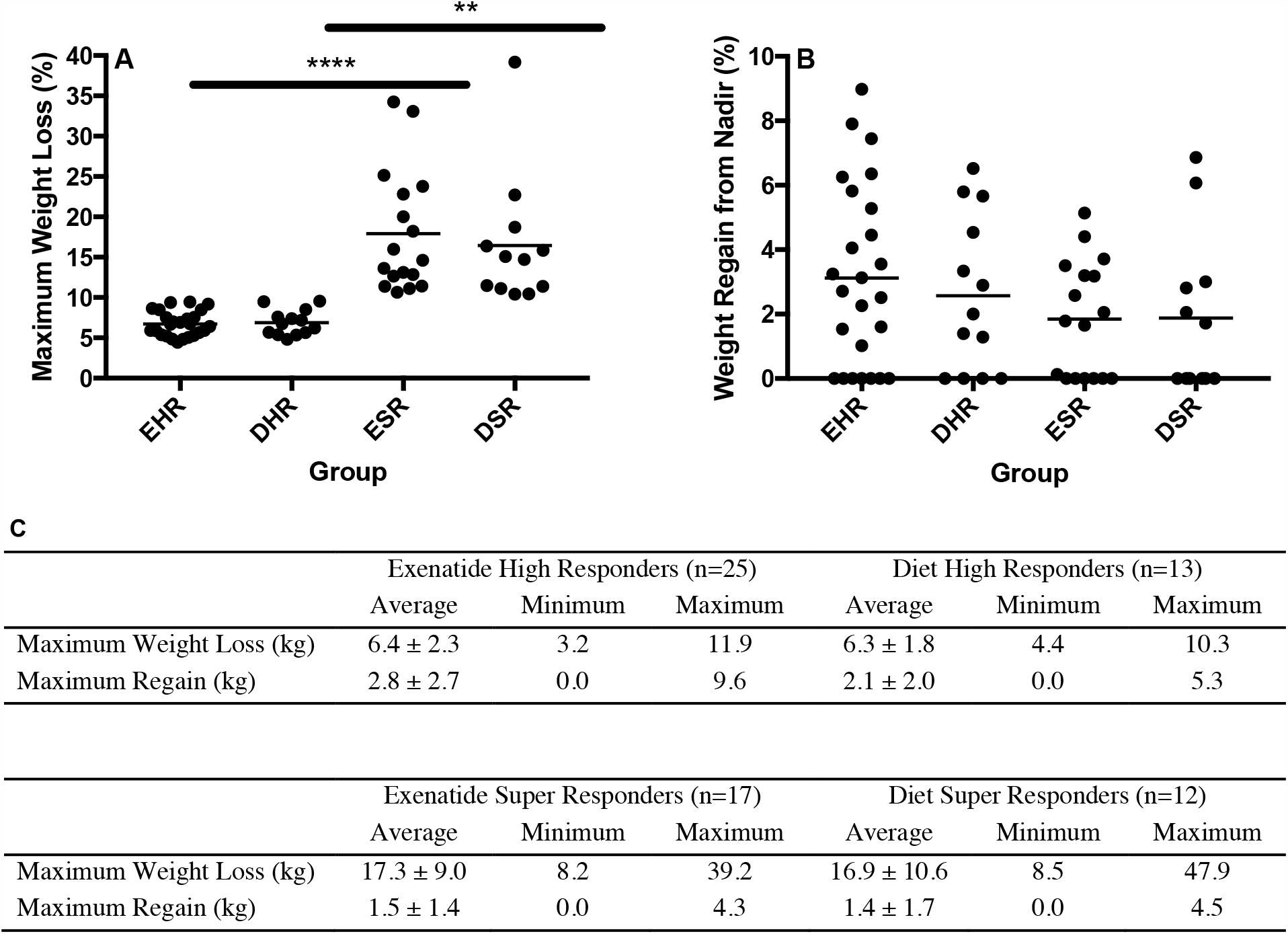
(A) Maximum weight loss (%) among high responders and super responders in both treatment groups **** = p<0.0005, ** = p<0.005 (B) Percentage weight regain from lowest weight achieved among high responders and super responders in both treatment groups. (C) Maximum weight loss and regain from lowest weight achieved in exenatide and diet high responder and super high responder groups. Values are mean +/- SD. EHR, exenatide high responder. ESR, exenatide super responder, DHR diet high responder, DSR diet super responder

Super high responders accounted for 23% of individuals treated with exenatide and 36% of those treated with diet; this difference was not significant (Figure 1A). There was similarly no significant difference in maximum percent weight loss between exenatide and diet super responder groups (p=0.62). In both groups, greater weight loss at 12 weeks predicted super responder status (exenatide *p=*0.0080, diet *p*=0.0098). Super high responders also demonstrated more than twice the total weight change and weight change at 3 months compared to high responders (Figure 2C).

### Metabolic Parameters

Both exenatide and diet high responders showed significant reductions in waist circumference, resting energy expenditure, and triglycerides (Table 2). Diet but not exenatide high responders also showed significant reductions in systolic blood pressure and percent body fat (Table 2). Both exenatide high and low responders showed a significant reduction in total cholesterol and LDL cholesterol while only diet high responders showed similar reductions in these parameters (Table 2).

### Predictors of Weight Loss and Super Responder Status

We evaluated several metabolic parameters as potential predictors of weight loss and of super high responder status.

Baseline age, weight, BMI, waist circumference, total cholesterol, and resting energy expenditure were all tested as variables that might predict high responder status using odds ratio estimates. None of these variables served as predictors for either treatment group (Table S2).

Age, weight, BMI, total cholesterol and waist circumference were also tested as predictors of super high responder status in both treatment groups. The distribution of these predictors by super high responder status shows that this group was slightly older and had higher total cholesterol levels and larger waist circumferences compared to high responders (Table S2).

### Metabolomics Analysis

In an exploratory analysis, we sought to identify metabolites that were associated with exenatide treatment response by leveraging a targeted LC-MS method that measures known metabolites. When comparing exenatide responders to non-responders, we found that exenatide responders had significantly lower baseline cysteine levels compared to exenatide non-responders (*p* <0.0001, Figure 4A). Next, we compared exenatide responders to diet responders and found higher baseline glucose, caffeine, and C6 carnitine (hexanoylcarnitine) levels in exenatide responders (*p* ≤0.04, Figure 4B). Finally, diet high responders exhibited increased niacinamide, anthranilic acid, and glycerol levels at baseline compared to diet low responders (p <0.04, Figure 5, Table S3), while Exenatide high responders had higher anandamide, sarcosine, X5 hydroxytryptophan, C18 carnitine, aminoisobutyric acid, serotonin, and N-acetyl-L-alanine (p <0.05, Figure 5, Table S3).

**Figure 4:**
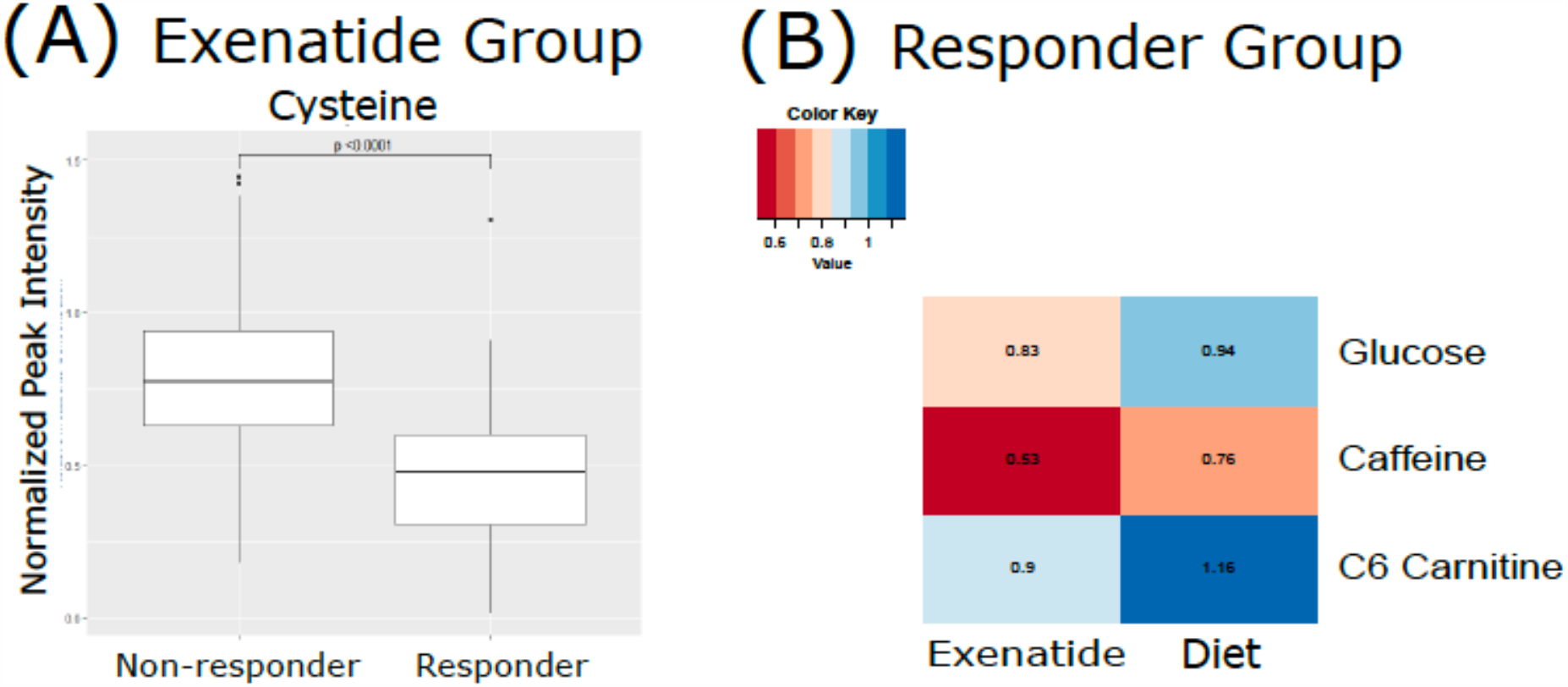
A) Mean normalized LC-MS peak intensity for cysteine, the only metabolite that was statistically different based on Wilcoxon Rank Sum test, between non-responder and responders in the Exenatide treated group. B: Heatmap depicting the differences in metabolite levels, as measured by LC-MS peak intensity, that were statistically different (p<0.05) by Wilcoxon Rank Sum test between responders in the Exenatide and Diet treated groups.

**Figure 5.**
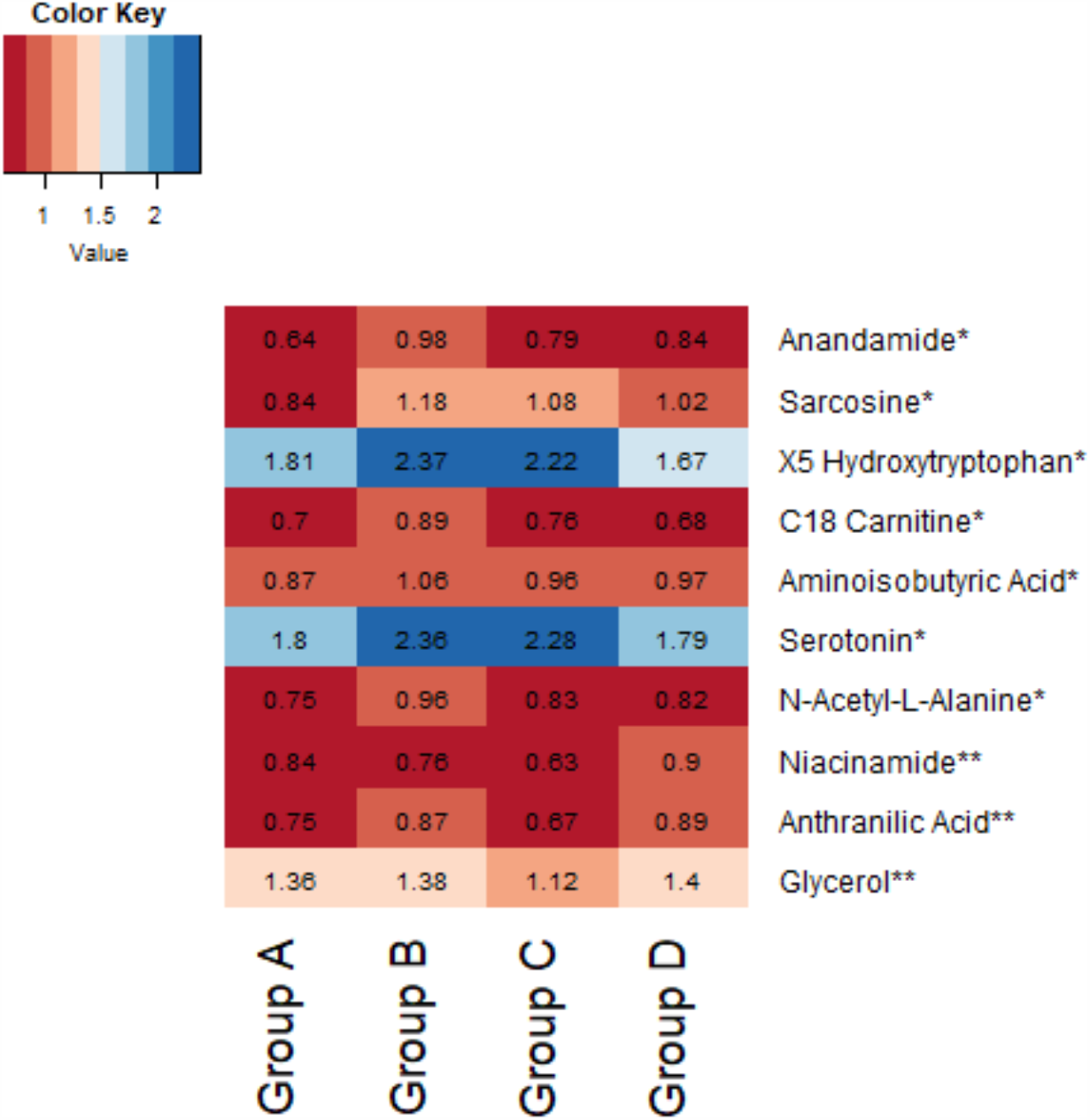
Heatmap demonstrating difference in baseline normalized peak intensities of metabolites between exenatide low responders (Group A, n = 22), exenatide high responders (Group B, n = 14), diet low responders (Group C, n = 11), and diet high responders (Group D, n = 12). *Metabolite peak intensities statistically significant between Group A and B based on Wilcoxon Rank Sum tests (p<0.05). **Metabolite peak intensities statistically significant between Group C and D based on Wilcoxon Rank Sum tests (p<0.05).

### Thermic Effect of Food

The thermic effect of food (TEF) was measured at baseline and after three months of either exenatide or diet treatment. At baseline, there were no differences between the groups (TEF 4.32 +/- 2.1 kcal/kg in the exenatide group and 4.49 +/- 2.0 kcal/kg in the diet group, Table 2). After 3 months of treatment, both exenatide high responders and low responders showed a decrease in the thermic effect of food (high responders – 0.96 kcal/kg, p=0.04, low responders – 1.55 kcal/kg, p=0.006, Table 2) while diet high and low responders showed no change in the thermic effect of food (high responders - 0.37 kcal/kg p=0.56, low responders +0.05 p=0.56, Table 2).

### Adverse Events

The most common adverse events for the study population as a whole were nausea (57%), decreased appetite (36%), and headache (26%). There was significantly more nausea among those randomized to exenatide compared to diet/placebo (70% vs 25%, p < 0.0001). Decreased appetite was also significantly more common in the exenatide versus diet/placebo group (41% compared to 24%, p =0.03). Headache was not significantly different between the two groups (27% in the exenatide group, 24% in the diet/placebo group, p=0.66). Among high responders, reports of decreased appetite were not significantly different between exenatide and diet/placebo groups (p=0.86). The incidence of nausea, decreased appetite, and headache decreased at each visit throughout the study in both treatment groups.

### Dropouts

Out of 249 individuals enrolled in the study, 182 were randomized to treatment and 67 were not randomized due to not randomized due based on disqualifying screening labs, withdrawal of consent, or lost to follow-up prior to randomization. The most common reasons subjects cited for dropping out were adverse events (20.3%) and time commitment (14.5%).

## Discussion

Weight loss can result in improvement or even reversal of metabolic sequelae of obesity^17^. GLP-1R agents are associated with significant long-term weight loss and are very rapidly gaining popularity for the treatment of obesity^11–14^ Clinicians’ experiences confirm large clinical studies showing a range of weight loss with these agents, however. Understanding the heterogeneity of weight loss response to specific pharmacotherapeutics, as well as hypocaloric diet, has important clinical and public health implications. The present study examines patterns and predictors of weight loss associated with twice daily exenatide treatment in overweight and obese women without diabetes.

In the present study, 56% of those treated with twice daily exenatide lost ≥ 5% body weight after 12 weeks of treatment compared to 76% of those treated with hypocaloric diet and matched placebo injections. This was not statistically significant, however a larger study might find a significant difference in weight loss when comparing exenatide to lifestyle modification. Importantly, in the present study, the group randomized to hypocaloric diet also injected a placebo twice daily before meals. These important behavioral interventions, specifically the pause before eating to self-administer a subcutaneous injection, may have affected food intake by making people more mindful about the meal they were about to eat, and more invested in the hypocaloric diet.

A subset of individuals whom we classified as super high responders demonstrated more robust weight loss, ≥ 10% of their body weight. Forty three percent of exenatide responders and 55% of diet high responders were super responders. As with high responders, this difference was not statistically significant, but presents the same trend toward a higher number of super high responders in the diet group. A variable weight loss response to exenatide treatment was apparent early on, with higher initial weight loss at 3 months predicting super high responder status. This finding is consistent with our previous study showing that the differential response to exenatide is evident early on during the treatment course^12^. Similar results have been seen with liraglutide^14^.

In addition to achieving similar average weight loss, both groups also demonstrated similar maximum weight loss, although the range of maximum weight loss was greater in the diet group. There was more weight regain among individuals in the exenatide group compared to the diet group, which could be explained by the treatment plateau that is seen with all medications prescribed for weight loss, and/or the lack of lifestyle change in the exenatide group. The diet/placebo group may have had less regain because these individuals made ongoing changes to food intake that were more were more durable than the effect of exenatide. Future studies might compare weight loss with exenatide treatment with and without diet and nutrition counseling in order to better isolate the pharmacotherapeutic effect of exenatide on body weight.

The thermic effect of food (TEF), which comprises approximately 8-10% of total daily energy expenditure, is typically unaffected by weight loss^18^. In this study, 12 weeks of exenatide treatment led to a significant decreased in TEF in both the high and low responder groups, while TEF was unchanged in the diet group. This suggests that the reduction in TEF is specifically attributable to exenatide, perhaps delayed gastric emptying specifically, rather than weight loss. Previous studies have similarly shown that treatment with GLP1R agonists leads to a decrease in TEF^19,20^. The compensatory metabolic mechanisms that favor weight loss despite the decrease in TEF among exenatide high responders remain unknown but are an important area of future research.

Exenatide treatment led to reductions in total and LDL cholesterol levels among both high and low weight loss responders, while these reductions were seen in only the diet high responder group. This agrees with a recent study showing that once-weekly exenatide leads to a decrease in total cholesterol^21^. This finding may be explained by the fact that even low responders to exenatide achieved modest weight loss, and/or a specific cholesterol lowering mechanism related to GLP1R agonism that is separate from weight loss. Interestingly, the present study did not show a significant change in triglycerides among those treated with exenatide, including high responders, although several human and rodent studies have reported significant decreases in triglycerides following treatment with GLP-1R agents^21,22^.

We did not identify any statistically significant predictors of weight loss, likely due to small sample size, however our data reveal interesting trends despite wide confidence intervals. The direction of the effect of baseline weight suggests that individuals with higher starting BMIs have a higher likelihood of reaching super responder status. Specifically, if baseline weight is 1kg higher, the odds of being super responder is increased by .7% (odds ratio of 1.007). Similarly, an increase of 1 year in age increases the odds of being a super responder by 1.2%, and every 1 unit increase in total cholesterol was associated with a 0.4% increase toward super responder status.

In exploratory analyses leveraging a targeted metabolomics platform, we found an association of lower baseline circulating cysteine concentrations with weight loss following exenatide treatment. Cysteine is an organosulfur compound that plays important roles in many enzymatic reactions in the body including redox reactions, metal coordination, and structural disulfide formation. It is also the precursor to the antioxidant glutathione. In epidemiology studies, total cysteine levels have been positively correlated with BMI and obesity^23,24^Moreover, modulating the levels of cysteine has been demonstrated to affect appetite and weight in rodent models^25,26^.

We also found a trend toward higher baseline levels of serotonin, aminoisobutyric acid, anandamide, and sarcosine in exenatide super responders. Anandamide is a lipid metabolite and neurotransmitter that plays a role in feeding behavior and obesity^26^. Aminoisobutyric acid is known to increase fatty acid oxidation and to decrease fat mass^27^. Notably, aminoisobutryric acid increases following exercise and induces browning of white fat^28^. In sum, the metabolite findings in our sub study point toward metabolite pathways that may impact biological pathways related to the weight loss, however future studies in a larger cohort are required to validate these findings. We acknowledge that a limitation of the metabolomics analysis is our small sample size which meant we were underpowered to definitively determine differences in metabolite concentration associations with treatment effect and our results could be biased by outliers.

The most common adverse events of nausea and decreased appetite were more prevalent in the exenatide group than the diet group, and more high responders to exenatide experienced nausea than low responders. Nausea itself may have contributed to the higher weight loss among exenatide responders, however as the study progressed, the incidence of side effects diminished in both exenatide groups.

In the field of obesity medicine, it is of increasing interest to identify predictors of pharmacologic success in order individualize an appropriate course of pharmacologic treatment. A recent study examined predictors of A1C reduction with exenatide treatment in individuals with type 2 diabetes and found that elevated initial A1C was the strongest predictor of magnitude of A1C reduction^29^. In our study, none of the metabolic variables we examined were identified as strong predictors of high responder status, but we did find that greater weight loss early in the treatment period predicted super high responder status. Future studies will need to examine additional predictor parameters including potential genetic factors that could contribute to this variable response.

This study has several limitations. All of the subjects in this study were relatively healthy women with overweight or obesity and without severe metabolic complications, thus the generalizability of these findings to a more heterogeneous population with obesity is limited. Future studies may include men and/or women with more poorly compensated obesity. Additionally, pharmacotherapies that are approved for weight loss are meant to be used only in combination with lifestyle modification, and in this study, we did not include lifestyle modification with exenatide treatment.

## Conclusion

The present study examined patterns and predictors of weight loss in overweight and obese women treated with exenatide or diet/placebo for at least 12 weeks. We found a highly variable response to exenatide within our study population, and we did not identify predictors of weight loss response. Our study suggests that while exenatide may have a few specific metabolic benefits, weight loss is similar to that achieved with a prescribed hypocaloric diet and placebo injections. Additionally, regain potential is higher with exenatide treatment.

## Data Availability

All data are housed in a HIPAA compliant database, RedCap, stored behind the firewall of the Beth Israel Deaconess Medical Center.

---

**Figure S1:**
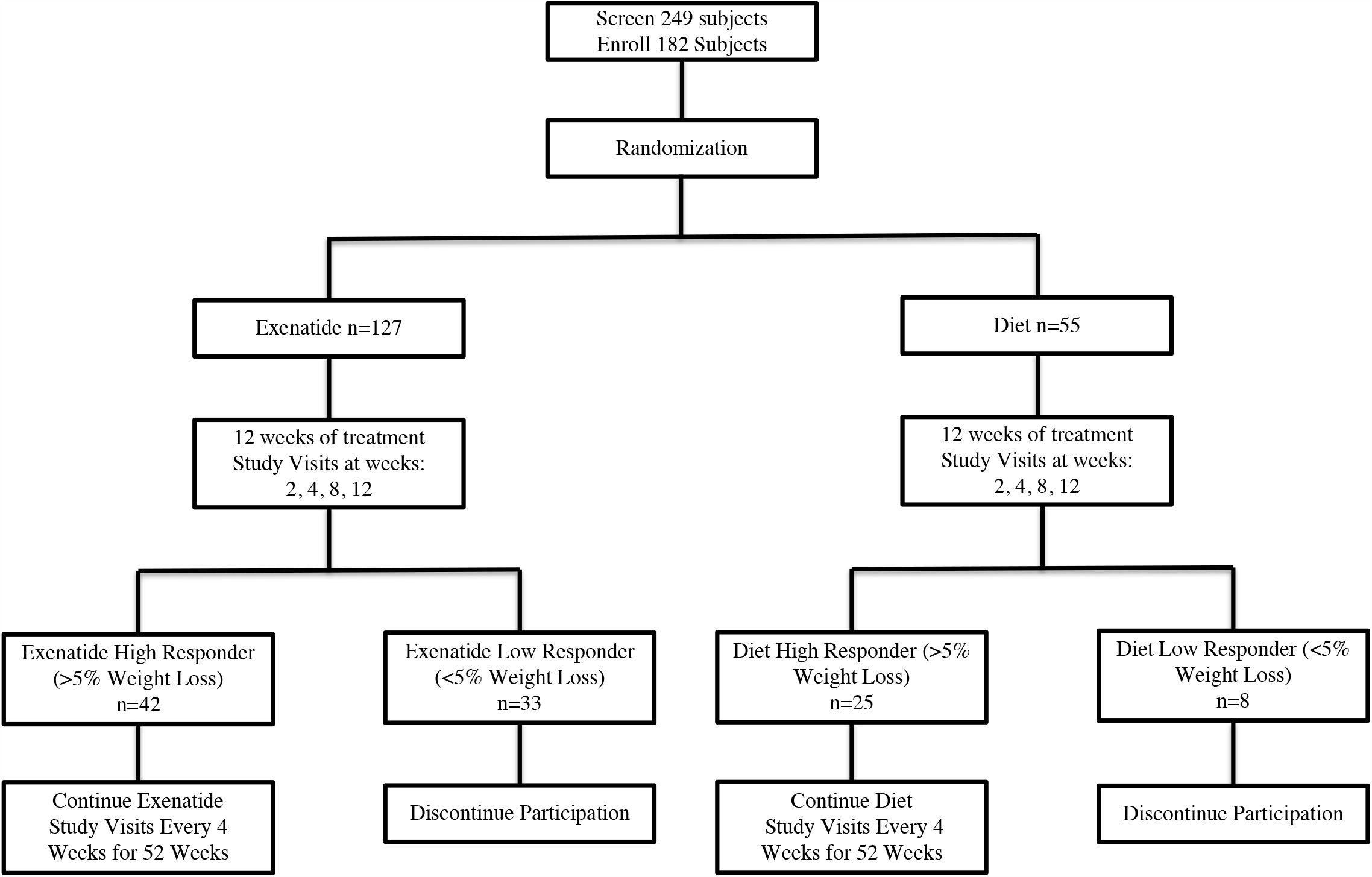
Study design.

**Table S1:** Baseline and week 12 characteristics of all responders (n=67)

|  | Week 0 | Week 12 |
| --- | --- | --- |
| Age (years) | 44.2 ± 12.4 | -- |
| BMI (kg/m<sup>2</sup>) | 35.5 ± 5.2 | 33.0 ± 4.6 |
| Weight (kg) | 94.2 ± 14.3 | 87.6 ± 12.9 |
| Systolic BP (mmHg) | 123.5 ± 12.4 | 118.5 ± 13.0 |
| Diastolic BP (mmHg) | 70.3 ± 9.3 | 71.6 ± 10.2 |
| Heart Rate (bpm) | 68.1 ± 10.5 | 68.1 ± 11.1 |
| Total Cholesterol (mg/dL) | 195.6 ± 3.9 | 178.7 ± 33.2 |
| HDL Cholesterol (mg/dL) | 60.5 ± 16.7 | 52.7 ± 14.6 |
| LDL Cholesterol (mg/dL) | 114.7 ± 3.6 | 108.0 ± 3.7 |
| Triglycerides (mg/dL) | 108.2 ± 56.5 | 89.8 ± 39.9 |
| Hemoglobin A1c (%) | 5.6 ± 0.3 | -- |
| Waist Circumference (cm) | 111.7 ± 10.9 | 104.1 ± 13.1 |
| Resting Energy Expenditure (kCal/d) | 1546.6 ± 212.5 | 1402.1 ± 162.6 |
| Percent Body Fat (%) | 43.7 ± 3.9 | 42.8 ± 4.4 |

**Table S2:** (A) Metabolic predictors of high responder status for both treatment groups (B) Metabolic predictors of super responder status for both treatment groups.

| **A** |  |  |
| --- | --- | --- |
|  | Odds Ratio | Confidence Interval |
| Age (years) | 1.03 | 0.97 – 1.09 |
| Weight (kg) | 0.98 | 0.98 – 1.20 |
| BMI (kg/m<sup>2</sup>) | 0.75 | 0.56 – 1.01 |
| Waist Circumference (cm) | 1.05 | 0.96 – 1.13 |
| Total Cholesterol (mg/dL) | 0.99 | 0.97 – 1.01 |
| Resting Energy Expenditure (kCal/d) | 1.16 | 0.94 – 1.42 |

| **B** |  |  |
| --- | --- | --- |
|  | Adjusted Odds Ratio | Confidence Interval |
| Age (years) | 1.01 | 0.98 – 1.05 |
| Weight (kg) | 1.007 | 0.98 – 1.04 |
| Waist Circumference (cm) | 2.00 | 0.84 – 4.83 |
| Total Cholesterol (mg/dL) | 1.00 | 0.99 – 1.02 |

**Table S3.** Differences in baseline metabolite levels in responders compared to super responders within treatment groups.

| Exenatide Treatment Group |  |  |  |
| --- | --- | --- | --- |
| Metabolite | Responder (n = 22) | High Responder (n =14) | P |
|  | Mean (SD) | Mean (SD) |  |
| Anandamide | 0.64 (0.20) | 0.98 (0.35) | 0.001 |
| Sarcosine | 0.84 (0.29) | 1.18 (0.27) | 0.002 |
| X5 Hydroxytryptophan | 1.81 (0.86) | 2.37 (0.77) | 0.01 |
| C18 Carnitine | 0.70 (0.25) | 0.89 (0.22) | 0.02 |
| Aminoisobutyric Acid | 0.87 (0.30) | 1.06 (0.21) | 0.02 |
| Serotonin | 1.80 (0.83) | 2.36 (0.77) | 0.02 |
| N-Acetyl-L-Alanine | 0.75 (0.26) | 0.96 (0.26) | 0.04 |
| Diet Treated Group |  |  |  |
| Metabolite | Responder (n = 11) | High Responder (n = 12) | P |
|  | Mean (SD) | Mean (SD) |  |
| Niacinamide | 0.63 (0.18) | 0.90 (0.25) | 0.02 |
| Anthranilic Acid | 0.67 (0.13) | 0.89 (0.27) | 0.02 |
| Glycerol | 1.12 (0.25) | 1.40 (0.32) | 0.02 |

Data is presented as mean (standard deviation) LC-MS spectra peak intensity after normalization to pooled plasma for each metabolite in Exenatide treated responders (n = 22) compared to Exenatide super responders (n = 14) and diet treated responders (n = 11) compared to placebo super responders (n = 12). P values were calculated using Wilcoxon rank sum on log transformed peak areas.

**Table S4:**
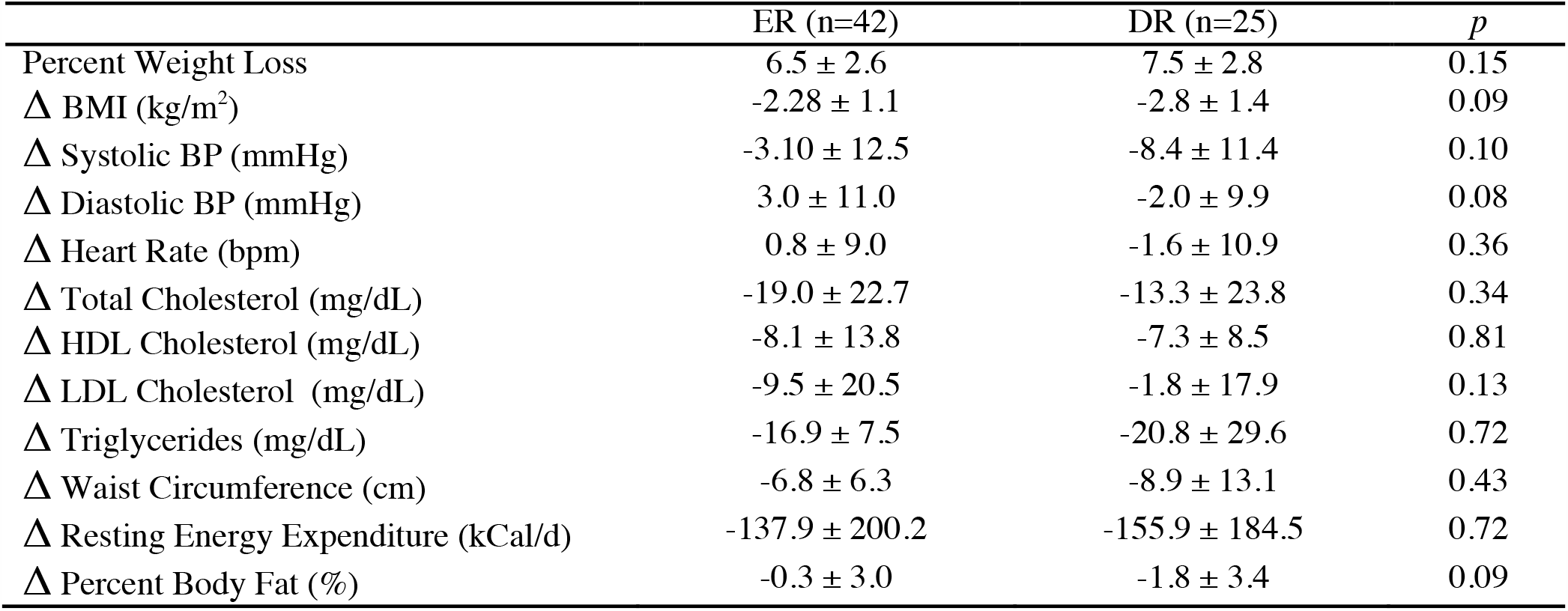
Aggregate data of responder characteristics (including both high responders and super responders) after twelve weeks of either exanetide or diet. Values are shown as mean ± SD. P values >0.05 were considered significant. ER exenatide responder, DR diet responder

## Notes

### Competing Interest Statement

The authors have declared no competing interest.

### Clinical Trial

NCT01590433

### Funding Statement

This study was an investigator initiated, industry funded study. Initial funding source was Amylin Pharmaceuticals, which was bought by Bristol Meyers Squibb and then Astra Zeneca. The sponsor is therefore referred to as Astra Zeneca.

### Author Declarations

Beth Israel Deaconess Medical Center Institutional Review Board

## References

1. Hales CM, Carroll MD, Fryar CD, Ogden CL. Prevalence of Obesity Among Adults and Youth: United States, 2015–2016 Key findings Data from the National Health and Nutrition Examination Survey. Natl Cent Heal Stat. 2017;1(288):1–8. https://www.cdc.gov/nchs/data/databriefs/db288.pdf.

2. Wadden TA. Eight-year weight losses with an intensive lifestyle intervention: The look AHEAD study. Obesity. 2014;22(1):5–13. doi:10.1002/oby.20662

3. Holzapfel C, Cresswell L, Ahern AL, et al. The challenge of a 2-year follow-up after intervention for weight loss in primary care. Int J Obes. 2014;38(6):806–811. doi:10.1038/ijo.2013.180

4. Curioni CC, Lourenço PM. Long-term weight loss after diet and exercise: A systematic review. Int J Obes. 2005;29(10):1168–1174. doi:10.1038/sj.ijo.0803015

5. Garvey T, Hurley DL, Mcmahon M, et al. Clinical Practice Guidelines for the Perioperative Nutritional. 2014;21(0 1):1–64. doi:10.1002/oby.20461.Clinical

6. Wharton S, Serodio KJ, Kuk JL, Sivapalan N, Craik A, Aarts M-A. Interest, views and perceived barriers to bariatric surgery in patients with morbid obesity. Clin Obes. 2016;6(2):154–160. doi:10.1111/cob.12131

7. Sarwer DB, Ritter S, Wadden TA, Spitzer JC, Vetter ML, Moore RH. Attitudes about the safety and efficacy of bariatric surgery among patients with type 2 diabetes and a body mass index of 30-40 kg/m2. Surg Obes Relat Dis. 2013;9(5):630–635. doi:10.1016/j.soard.2012.10.007

8. Doyle ME, Egan JM. Mechanisms of action of glucagon-like peptide 1 in the pancreas. Pharmacol Ther. 2007;113(3):546–593. doi:10.1016/j.pharmthera.2006.11.007

9. Pi-Sunyer X, Astrup A, Fujioka K, et al. A Randomized, Controlled Trial of 3.0 mg of Liraglutide in Weight Management. N Engl J Med. 2015;373(1):11–22. doi:10.1056/NEJMoa1411892

10. Kim SH, Abbasi F, Lamendola C, et al. Benefits of liraglutide treatment in overweight and obese older individuals with prediabetes. Diabetes Care. 2013;36(10):3276–3282. doi:10.2337/dc13-0354

11. Nuffer WA, Trujillo JM. Liraglutide: A New Option for the Treatment of Obesity. Pharmacotherapy. 2015;35(10):926–934. doi:10.1002/phar.1639

12. Dushay J, Gao C, Gopalakrishnan GS, et al. Short-term exenatide treatment leads to significant weight loss in a subset of obese women without diabetes. Diabetes Care. 2012;35(1):4–11. doi:10.2337/dc11-0931

13. O’Neil P, Birkenfeld A, McGowan B, et al. Efficacy and safety of semaglutide compared with liraglutide and placebo for weight loss in patients with obesity: a randomised, double-blind, placebo and active controlled, dose-ranging, phase 2 trial. Lancet. 2018;392. doi:10.1016/S0140-6736(18)31773-2

14. Fujioka K, O’Neil PM, Davies M, et al. Early Weight Loss with Liraglutide 3.0 mg Predicts 1-Year Weight Loss and is Associated with Improvements in Clinical Markers. Obesity. 2016;24(11):2278–2288. doi:10.1002/oby.21629

15. Kimberly WT, O’Sullivan JF, Nath AK, et al. Metabolite profiling identifies anandamide as a biomarker of nonalcoholic steatohepatitis. JCI Insight. 2017;2(9). doi:10.1172/jci.insight.92989

16. Wang TJ, Larson MG, Vasan RS, et al. Metabolite profiles and the risk of developing diabetes. Nat Med. 2011;17(4):448–453. doi:10.1038/nm.2307

17. Lean MEJ, Leslie WS, Barnes AC, et al. Primary care-led weight management for remission of type 2 diabetes (DiRECT): an open-label, cluster-randomised trial. Lancet. 2018;391(10120):541–551. doi:10.1016/S0140-6736(17)33102-1

18. Rosenbaum M, Leibel RL. Adaptive Thermogenesis in. 2013;34(0 1):1–15. doi:10.1038/ijo.2010.184.Adaptive

19. Maciel MG, Beserra BTS, Oliveira FCB, et al. The effect of glucagon-like peptide 1 and glucagon-like peptide 1 receptor agonists on energy expenditure: A systematic review and meta-analysis. Diabetes Res Clin Pract. 2018;142:222–235. doi:10.1016/j.diabres.2018.05.034

20. Holst J, Ersbøll A, Astrup A, Flint A, Raben A. The effect of physiological levels of glucagon-like peptide-1 on appetite, gastric emptying, energy and substrate metabolism in obesity. Int J Obes. 2002;25(6):781–792. doi:10.1038/sj.ijo.0801627

21. Davies MJ, Bergenstal R, Bode B, et al. Efficacy of liraglutide for weight loss among patients with type 2 diabetes: The SCALE diabetes randomized clinical trial. JAMA - J Am Med Assoc. 2015;314(7):687–699. doi:10.1001/jama.2015.9676

22. Cummings BP, Stanhope KL, Graham JL, et al. Chronic administration of the glucagon-like peptide-1 analog, liraglutide, delays the onset of diabetes and lowers triglycerides in UCD-T2DM rats. Diabetes. 2010;59(10):2653–2661. doi:10.2337/db09-1564

23. Elshorbagy AK, Smith AD, Kozich V, Refsum H. Cysteine and Obesity. Obesity. 2012;20(3):473–481. doi:10.1038/oby.2011.93

24. Elshorbagy AK, Kozich V, Smith AD, Refsum H. Cysteine and obesity: consistency of the evidence across epidemiologic, animal and cellular studies. Curr Opin Clin Nutr Metab Care. 2012;15(1). https://journals.lww.com/co-clinicalnutrition/Fulltext/2012/01000/Cysteine_and_obesityconsistency_of_the_evidence.9.aspx.

25. Elshorbagy AK, Valdivia-Garcia M, Mattocks DAL, et al. Cysteine supplementation reverses methionine restriction effects on rat adiposity: significance of stearoyl-coenzyme A desaturase. J Lipid Res. 2011;52(1):104–112. doi:10.1194/jlr.M010215

26. Gaetani S, Kaye WH, Cuomo V, Piomelli D. Role of endocannabinoids and their analogues in obesity and eating disorders. Eat Weight Disord EWD. 2008;13(3):e42–8.

27. Begriche K, Massart J, Abbey-Toby A, Igoudjil A, Lettéron P, Fromenty B. β-Aminoisobutyric Acid Prevents Diet-induced Obesity in Mice With Partial Leptin Deficiency. Obesity. 2008;16(9):2053–2067. doi:10.1038/oby.2008.337

28. Roberts LD, Boström P, O’Sullivan JF, et al. β-Aminoisobutyric Acid Induces Browning of White Fat and Hepatic β-Oxidation and Is Inversely Correlated with Cardiometabolic Risk Factors. Cell Metab. 2014;19(1):96–108. doi:10.1016/j.cmet.2013.12.003

29. Khan M, Ouyang J, Perkins K, Nair S, Joseph F. Determining Predictors of Early Response to Exenatide in Patients with Type 2 Diabetes Mellitus. J Diabetes Res. 2015;2015:1–9. doi:10.1155/2015/162718

